# Aberrant oscillatory activity in Neurofibromatosis Type 1: An EEG study of resting state and working memory

**DOI:** 10.1101/2022.08.24.22279165

**Authors:** Samantha J. Booth, Shruti Garg, Laura J. E. Brown, Jonathan Green, Gorana Pobric, Jason R. Taylor

**Affiliations:** Division of Human Communication, Development, and Hearing, School of Health Sciences, Faculty of Biology, Medicine, and Health, University of Manchester, Manchester Academic Health Science Centre, England, UK; Division of Psychology and Mental Health, School of Health Sciences, Faculty of Biology, Medicine, and Health, University of Manchester, Manchester Academic Health Science Centre, England, UK; Child & Adolescent Mental Health Services, Royal Manchester Children’s Hospital, Central Manchester University Hospitals NHS Foundation Trust, Manchester Academic Health Science Centre, England, UK

**Keywords:** Electroencephalography (EEG), neurofibromatosis type 1 (NF1), oscillations, oscillatory power, phase coherence, working memory

## Abstract

**Background:** Neurofibromatosis Type 1 (NF1) is a genetic neurodevelopmental disorder commonly associated with impaired cognitive function. Despite the well-explored functional roles of neural oscillations in neurotypical populations, only a limited number of studies have investigated oscillatory activity in the NF1 population.

**Methods:** We compared oscillatory spectral power and theta phase coherence in a paediatric sample with NF1 (N=16; mean age: 13.03 years; female: n=7) to an age/sex-matched typically-developing control group (N=16; mean age: 13.34 years; female: n=7) using electroencephalography measured during rest and during working memory task performance.

**Results:** Relative to typically-developing children, the NF1 group displayed higher resting state slow wave power and a lower peak alpha frequency. Moreover, higher theta power and frontoparietal theta phase coherence were observed in the NF1 group during working memory task performance, but these differences disappeared when controlling for baseline (resting state) activity.

**Conclusions:** Overall, results suggest that NF1 is characterised by aberrant resting state oscillatory activity that may contribute towards the cognitive impairments experienced in this population.

**Trial registration:** ClinicalTrials.gov identifier: NCT03310996 (first posted: October 16 2017).

## 1. Background

Neurofibromatosis Type 1 (NF1) is an autosomal-dominant neurodevelopmental disorder, present in around 1 in 2700 births [1]. Although there is great inter-individual variability in its clinical manifestations, core somatic symptoms include dermal neurofibromas and pigmentary lesions [2]. In addition to somatic symptoms, social and behavioural difficulties are common, with around 50% of children with NF1 meeting the diagnostic criteria for attention-deficit hyperactivity disorder (ADHD) and around 25% for autism spectrum disorder (ASD) [3]. Moreover, cognitive impairments, including working memory deficits, are prevalent [4] and substantially impair academic achievement [5] and impact negatively on quality of life [6].

There remains a need to better understand the relationship between cortical function and cognitive deficits in NF1 [7] to help provide target(s) for pharmacological and non-pharmacological interventions (e.g., non-invasive brain stimulation [8]; neurofeedback: [9]), which in turn may improve treatment outcomes and academic trajectories. Existing neuroimaging research has related cognitive deficits in NF1 to brain function using functional magnetic resonance imaging (fMRI). Studies suggest aberrant activity compared to typically-developing controls [10]. For example, increased functional connectivity between the ventral anterior cingulate cortex and the insular cortex during rest may contribute to impaired cognitive control [11]. Aberrant activity has also been observed during cognitive task performance, including reduced task-related activity in key frontal and parietal regions during working memory tasks [12,13]. Only a limited number of studies have used M/EEG to investigate the neural correlates of cognitive impairments in NF1. Abnormalities in Event Related Potential (ERP) components relative to typically-developing controls have been reported [14,15,16,17]. For instance, reduced P1 amplitude has been observed, suggesting aberrant early visual processing [14,18]. Additionally, reduced P3a amplitude has been observed during a go/no-go task and is hypothesised to reflect impaired inhibition [18]. Furthermore, topographic differences in P3b amplitude and a shorter P3b latency relative to controls have been found during working memory task performance, which the authors suggest may contribute to the cognitive deficits seen in NF1 [16].

The high temporal resolution of M/EEG also enables the study of brain oscillations. Oscillations can be seen as a ‘primary’ or a more direct measure of brain activity relative to ERPs, therefore providing an important window into understanding cognitive processes [19]. As such, investigating oscillations in NF1 can provide additional insights to those gained from ERP methods. Oscillatory measures are particularly well-suited to the investigation of protracted processes, such as those required for working memory – a cognitive ability that is impaired in NF1 [16]. In healthy adults, increased (mid-frontal) theta power is observed during working memory maintenance and is hypothesised to maintain the temporal relationship between items held in working memory [20, 21]. Moreover, increased theta phase coherence (i.e., the consistency of phase values between brain regions [22]) between frontal and parietal-temporal regions is observed during working memory maintenance and is thought to facilitate integration of information between these key regions of the working memory network [23,24]. The literature exploring working memory related (WM-related) theta oscillations in typically-developing children is sparse [25], though there is some evidence to suggest that increased theta band activity occurs during working memory maintenance, like in adulthood [26,27].

Studies in neurodevelopmental disorders that experience overlapping cognitive impairments with NF1 (e.g., ADHD/ASD [14]) have reported differences in oscillatory activity during working memory performance relative to typically-developing controls [27,28,29,30,31]. For instance, Jang et al. [28] found significantly reduced theta power in ADHD relative to controls. Moreover, Yuk et al. [31] observed reduced phase coherence in ASD between key nodes of the working memory network (i.e., frontal and parietal regions) which has been hypothesised to reflect difficulty integrating information. Given that aberrant activity in specific frequency bands exhibits considerable overlap across neurodevelopmental disorders [32] we might also observe abnormalities in WM-related oscillatory activity in the NF1 population.

At present there are only two existing studies investigating M/EEG oscillatory correlates of cognitive impairments in NF1 [17,33]. These studies did not investigate activity during working memory task performance, but instead during rest and during visual processing [17] and covert attention tasks [33]. Ribeiro et al. [17] observed higher resting state theta power in NF1 relative to typically-developing controls, typical alpha reactivity (i.e., higher alpha power during eyes closed relative to eyes open resting state [34]), and enhanced alpha power during visual processing that may provide a neural marker of attentional deficits in this population. Moreover, Silva et al. [33] found elevated alpha desynchronisation during a covert attention task that may reflect a compensatory mechanism to keep performance at normal levels. Exploration of oscillatory activity in other cognitive domains impaired in NF1, such as working memory, is lacking. Investigating oscillatory activity during working memory is important as working memory underpins and shares common neural correlates with other cognitive functions important to everyday functioning, such as learning [35] and attention [36].

With this in mind, the current study compared EEG power and theta phase coherence in adolescents with NF1 to an age/sex-matched typically-developing control group using EEG measured during rest and during a working memory task. Consistent with previous work [17], we hypothesised higher resting state spectral power in NF1, but normal power reactivity during eyes open versus eyes closed resting state conditions. Our analysis of oscillatory activity during working memory was largely exploratory; however, we predicted aberrant theta power and frontoparietal theta phase coherence given previous WM-related EEG studies in other neurodevelopmental disorders [29,31]. Finally, the association between EEG measures and age, overall cognitive function, and working memory performance were also explored.

## 2. Methods

The current study is an extension of the analysis presented in Pobric et al. [16]. Specifically, we conducted oscillatory analyses on the same participants, using the same EEG resting state and n-back data, and used some (see Section 2.2) of the same behavioural measures described in Pobric et al. [16].

### 2.1 Participants

Thirty-two participants completed this study^1^. Participants were adolescents with NF1 (n=16) and age and sex-matched controls (n=16). Participants were required to meet each of the eligibility criteria in **Table 1**. Parents/guardians gave oral and written consent, and adolescents assent (where developmentally appropriate), prior to participation.

**Table 1.**
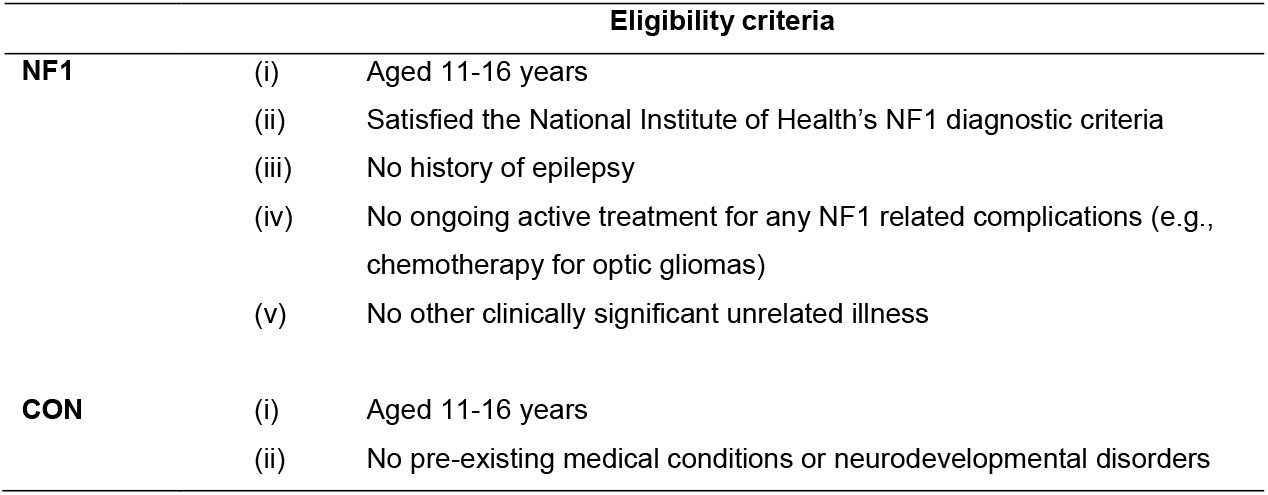
Eligibility criteria.

| Eligibility criteria |  |  |
| --- | --- | --- |
| NF1 | (i) | Aged 11-16 years |
|  | (ii) | Satisfied the National Institute of Health's NF1 diagnostic criteria |
|  | (iii) | No history of epilepsy |
|  | (iv) | No ongoing active treatment for any NF1 related complications (e.g., chemotherapy for optic gliomas) |
|  | (v) | No other clinically significant unrelated illness |
| CON | (i) | Aged 11-16 years |
|  | (ii) | No pre-existing medical conditions or neurodevelopmental disorders |

<sup>1</sup> See Section 4.5 for a consideration of power.
<sup>2</sup> This remained the case for the sub-group of participants included in the resting state (CON/NF1: n=16/14) and task-specific (CON/NF1: n=16/13) analyses (**Additional file 1**).

The NF1 sample were recruited through the Manchester Centre for Genomic Medicine, Neurofibromatosis charities, social media platforms, and newsletters, and were adolescents who satisfied the National Institute of Health’s (1988) diagnostic criteria for NF1 [37]. The control sample (CON) were age and sex-matched at group level and recruited via institutional newsletter advertisements and contacting local schools. Demographic information of the sample is reported in **Table 2**. There were no significant differences between groups in age (*t*_(30)_ = 0.540, *p*=.593) or sex (χ2 = 0.00, *p*=1.00)^2^.

**Table 2.** Participant demographics.

| Demographic | Group |  |
| --- | --- | --- |
|  | NF1 | CON |
| Age (M±SD (range)) | 13.03±1.66 (11.33-16.92) years | 13.34±1.61 (11.25-16.58) years |
| Sex (male/female) | 9/7 | 9/7 |
| NF1 mutation (inherited/de novo) | 7/9 | N/A |
| Medication | Melatonin (n=2) | N/A |
|  | Methylphenidate (n=4) |  |
| Pre-existing clinical diagnoses | ADHD and ASD (n=3) | N/A |
|  | ADHD (n=1) |  |
|  | ASD (n=2) |  |

### 2.2 Procedure

This study received ethical approval from the Greater Manchester West Research Ethics Committee (17/NW/0364) and was conducted in accordance with the Declaration of Helsinki. During the study visit, participants and their parents/guardians were first familiarised with the EEG equipment and study procedures. Subsequently, a battery of behavioural and cognitive assessments was administered, including parent-rated and cognitive measures that tapped into: overall cognitive function, inattention, hyperactivity, communication, daily living skills, socialisation, short-term memory, working memory, sustained attention, and attentional switching, followed by EEG. This paper focuses on parent-reported^3^ Adaptive Behaviour Composite (ABC) scores on the Vineland Adaptive Behaviour Scale (VABS-III) [38], performance on an adaptive auditory n-back task [16], and performance on a non-adaptive visual n-back task [16] performed during EEG (see Pobric et al. [16] for details of the other tasks performed that are not reported here).

VABS-II measures daily living skills, socialisation, and communication, with ABC scores reflecting standardised age equivalent overall cognitive functioning [38]. Performance on the adaptive auditory and non-adaptive visual n-back tasks measure working memory. Each trial of the n-back task began with a fixation cross (+) presented in the centre of the screen (adaptive auditory n-back: 2500ms; non-adaptive visual n-back: 2000ms, +/- random jitter up to 100ms in 17ms steps). This was followed by a single uppercase English consonant (C, G, H, K, P, Q, T, or W) presented aurally (auditory n-back: 1000ms) or visually in the centre of the screen (non-adaptive visual n-back: 500ms). Participants were instructed to respond as quickly and accurately as possible with a mouse-click whenever the current stimulus was the same as the one presented ‘n’ steps back in the sequence. No responses were required for non-targets. The auditory n-back was adaptive, such that after each block of 20 trials, the difficulty level of the next block was adjusted based on the participant’s performance to ensure participants were always training at the top of their ability (see Pobric et al. [16] for further task details). In contrast, the visual n-back performed during EEG recording was not designed to push participants’ ability to their limit. Instead, it was developed to provide a sufficient number of trials to permit investigation of electrophysiological differences between the groups during working memory performance. In this non-adaptive task, four fixed-order blocks were presented: 1-back, 2-back, 2-back, and 1-back, with self-paced breaks in between to reduce fatigue. In each block there were 100 trials, 25 of which were target trials (i.e., the same letter as ‘n’ screens back). As existing studies report a load-related increase in power during working memory maintenance [39], two load levels (‘n’ = 1 and 2) were included to permit investigation of load-dependent effects on EEG measures.

### 2.3 EEG acquisition

EEG data were recorded using an ActiveTwo system (BioSemi, Amsterdam, Netherlands) with 64 EEG channels in standard 10-10 system locations plus HEOG, VEOG, and mastoids, with a sampling rate of 512Hz. During the recording, participants were asked to remain still, in a comfortable/relaxed position, and to minimise eye-movements and blinking where possible. Recording started with 2.5 minutes of eyes open and 2.5 minutes of eyes closed resting state, in which participants were asked to simply relax and not think of anything in particular. This was followed by recording during the visual n-back task.

### 2.4 EEG analysis

MATLAB (2019a) and SPM12 (version 7771) [40] were used to conduct data analyses. Custom functions [41,42] calling several functions from EEGLAB (version 13.6.5b/v2020.0) [43] and FieldTrip [44] were used.

#### 2.4.1 Common pre-processing

Continuous EEG data were re-referenced to averaged mastoids, high-pass filtered (0.1Hz), down sampled (256Hz), low-pass filtered (resting state: 200Hz; task-related: 120Hz), and notch-filtered (48-52Hz), before epoching (resting state: arbitrary 1900ms (baseline correction: 0-1900ms, i.e., mean-centring); task-related: 0-1900ms relative to stimulus onset). The eyes open and eyes closed data were then concatenated (i.e., combined into the same file).

Independent Component Analysis (ICA) was used to project blink and eye-movement signals out of the data^4^. Channels containing noise unrelated to blinks (characterised by large positive deflections) or eye-movements (characterised by square-wave deflections) were temporarily omitted (channel TP7 was persistently bad and omitted from ICA for all participants). Thirty-two components were extracted from EEG channel data only. ICA components with uniquely high temporal correlations with VEOG and HEOG, and/or uniquely high spatial correlations with the blink topography, were identified using custom code [41] and following the procedure described in Pobric et al. [16]. The resulting weight matrix (less the artefact components) was applied to the epoched data using SPM12’s ‘montage’ function.

Baseline correction was then re-applied on the ICA-cleaned data. Epochs were rejected as noisy if they contained signal that exceeded a threshold (resting state: 200µV; task-related: 120µV; higher threshold for resting state data due to higher alpha power during eyes closed resting state). A channel was declared ‘bad’ if the threshold was exceeded on >20% of trials, and epoch rejection was re-run ignoring any bad channels. To reconstruct these noisy channels, a channel-weight interpolation matrix was created using FieldTrip’s ‘channelrepair’ function and applied to the epoched data using SPM12’s ‘montage’ function. EEG data were then re-referenced to the common average reference. The mean number of components removed, channels interpolated, and trials remaining can be seen in **Table 3**.

**Table 3.** Number of components removed, channels interpolated, and trials remaining.

|  | No. of components removed |  | No. of channels interpolated |  | No. of trials remaining<sup>5</sup> |  |  |  |
| --- | --- | --- | --- | --- | --- | --- | --- | --- |
|  | M±SD | Min-Max | M±SD | Min-Max | Open/1-back |  | Closed/2-back |  |
|  |  |  |  |  | M±SD | Min-Max | M±SD | Min-Max |
| *Resting-state* |  |  |  |  |  |  |  |  |
| CON | 1.94±0.44 | 1-3 | 1.00±0.00 | 1-1 | 75.13±11.63 | 48-105 | 71.19±6.39 | 52-77 |
| NF1 | 1.86±0.36 | 1-2 | 1.00±0.00 | 1-1 | 61.43±18.34 | 22-76 | 63.64±15.17 | 32-77 |
| *Task-related* |  |  |  |  |  |  |  |  |
| CON | 1.69±0.70 | 1-3 | 1.50±1.21 | 1-5 | 130.55±50.00 | 46-194 | 111.88±47.22 | 38-194 |
| NF1 | 1.73±0.59 | 1-3 | 1.73±1.62 | 1-6 | 132.33±46.24 | 28-188 | 113.13±53.10 | 38-195 |

Abbreviations: M: mean, SD: standard deviation, Min: minimum, Max: maximum.

#### 2.4.2 Resting state analysis

To be eligible for inclusion participants were required to have a minimum of 15 valid epochs remaining in each condition (open/closed) after artefact-contaminated trials were removed.

Two participants were excluded from the NF1 group, one due to having fewer than 15 valid trials in the eyes open condition and the other due to low quality data (i.e., all channels automatically marked ‘bad’ during the artefact detection routine). The sample used for the resting state analyses therefore comprised 30 participants (n=16 CON; n=14 NF1).

#### 2.4.3 Task-related analysis

Trials with incorrect responses were excluded. Subsequent analyses were conducted on target and non-target trial data without distinguishing between these conditions, since n-back performance requires maintenance of information during both trial types, particularly in the late post-ERP time window (described below). For the same reason, the number of non-target trials was not decimated to match the number of target trials (which was done in the P300 analysis presented Pobric et al., [16]).

To be eligible for inclusion participants were required to have a minimum of 15 valid epochs remaining in each load (1-/2-back) after artefact-contaminated trials and incorrect trials were removed. One participant from the NF1 group was excluded owing to having fewer than 15 valid trials in the 2-back load level (this was a different participant to the two resting state exclusions). The sample size used for the task-related analyses therefore comprised 31 participants (n=16 CON; n=15 NF1).

#### 2.4.4 Spectral power

For estimation of task-related power the time-window of interest was 900-1900ms post-stimulus onset (i.e., during the fixation cross of the next trial). This time-window was chosen as existing studies investigating WM-related oscillatory activity typically use the maintenance period of the working memory task as the time-window of interest as increased oscillatory activity is observed during this period [20,24,31,46]. We followed the previous literature’s definition of the maintenance period as the time following a response to the stimuli, determined using the average (or median) response time on the given task [31]. In the current study, the average response time over 1-/2-back blocks was 627±124ms (median: 601ms). However, to ensure that the majority of participants had responded, 900ms was chosen as the beginning of the time-window. The time window ended at 1900ms to provide a sufficient number of samples for power estimation. For consistency, the same time-window (in the arbitrary epoch), and therefore number of samples, was used for the estimation of resting state power.

For each EEG channel and epoch (resting state: eyes open and eyes closed; task-related: 1-back and 2-back), a Fast Fourier Transform with a Hanning window and a frequency resolution of 1Hz was used to extract frequency spectra collapsed over time (900-1900ms). The resulting power values were then log-transformed before averaging spectra over epochs. For exploratory analysis, average log-transformed power over all EEG channels was computed in canonical frequency bands: delta: 1-3Hz; theta: 4-7Hz; alpha: 8-11Hz; beta: 12-29Hz; low-gamma: 30-47Hz, and high-gamma: 53-100Hz. Additionally, as the literature consistently reports increased mid-frontal theta power during working memory maintenance [20], we performed targeted analysis of task-related mid-frontal theta (4-7Hz) power. To achieve this, log-transformed power was averaged over channels Fz, F1, and F2 to create a mid-frontal region of interest prior to statistical analysis. We measured absolute power (i.e., power in one frequency band, independent of activity in other frequency bands), as opposed to relative power (i.e., power in one frequency band divided by the amount of activity in all frequency bands) to avoid the potential confound that any abnormalities in one frequency band may affect the relative power of other frequency bands — a particular concern in neurodevelopmental disorder studies [47].

#### 2.4.5 Peak alpha frequency

Differences in peak alpha frequency (PAF) between groups were investigated. PAF was defined as the frequency with the maximum power in a loose alpha range (6.5-13.5Hz) at channel Pz. Pz was chosen as alpha power is typically high at this channel [48]. For analysis of PAF, for each EEG channel and epoch (eyes open/closed), a Fast Fourier Transform with a Hanning window and a frequency resolution of 0.25Hz was used to extract frequency spectra collapsed over time (arbitrary 1900ms epoch). First, each individual’s 1D spectrum was adjusted to reduce 1/f noise as this flattens the spectrum and causes the alpha peak to ‘pop out’ [49]. This was achieved by fitting a second-order polynomial to the log-transformed frequencies (omitting alpha and notch-filter frequencies), and the difference between the spectrum and this model was computed. The resulting spectrum was smoothed with a Gaussian kernel to remove spurious peaks. Next, four adjustments were made based on visual inspection of each participant’s spectrum: Two CON and one NF1 participant’s had maxima that fell on the ascending slope of the beta peak (eyes open: 12.25Hz, 13.25Hz, and 12.75Hz; eyes closed: 13.25Hz, 12.75Hz), and these were adjusted to small visible alpha peaks (eyes open: 11.50Hz, 10.25Hz, and 10.75Hz; eyes closed: 11.75 Hz, 11.75Hz) that our algorithm had missed; and one NF1 participant’s maximum fell on the descending delta slope (eyes open and closed: 6.5Hz) and were adjusted to small visible alpha peaks (eyes open: 7Hz, eyes closed: 7.5Hz) missed by the algorithm.

#### 2.4.6 Theta phase coherence

Prior to estimating phase coherence, the task-related data were spatially filtered using the Surface Laplacian, implemented using the laplacian_perrinX function in MATLAB [50]. The Surface Laplacian reduces the influence of volume conduction, which is particularly important given the electrode-level connectivity analysis performed [51]. We investigated theta phase coherence in the frontoparietal network **(Figure 1)**. The mid-frontal region acted as a seed region and coherence was estimated between this region and left-parietal, mid-parietal, and right-parietal regions [52]. Each region comprised of a set of electrodes: mid-frontal (F1/Fz/F2), left-parietal (P3/P5/P7), mid-parietal (P1/Pz/P2), and right-parietal (P4/P6/P8). Coherence was estimated between each possible mid-frontal – parietal connection (i.e., 27 channel pairs), before averaging coherence over electrode sets, resulting in three coherence estimates: mid-frontal to (1) left-parietal (ML), (2) mid-parietal (MM), and (3) right-parietal (MR).

**Fig. 1.**
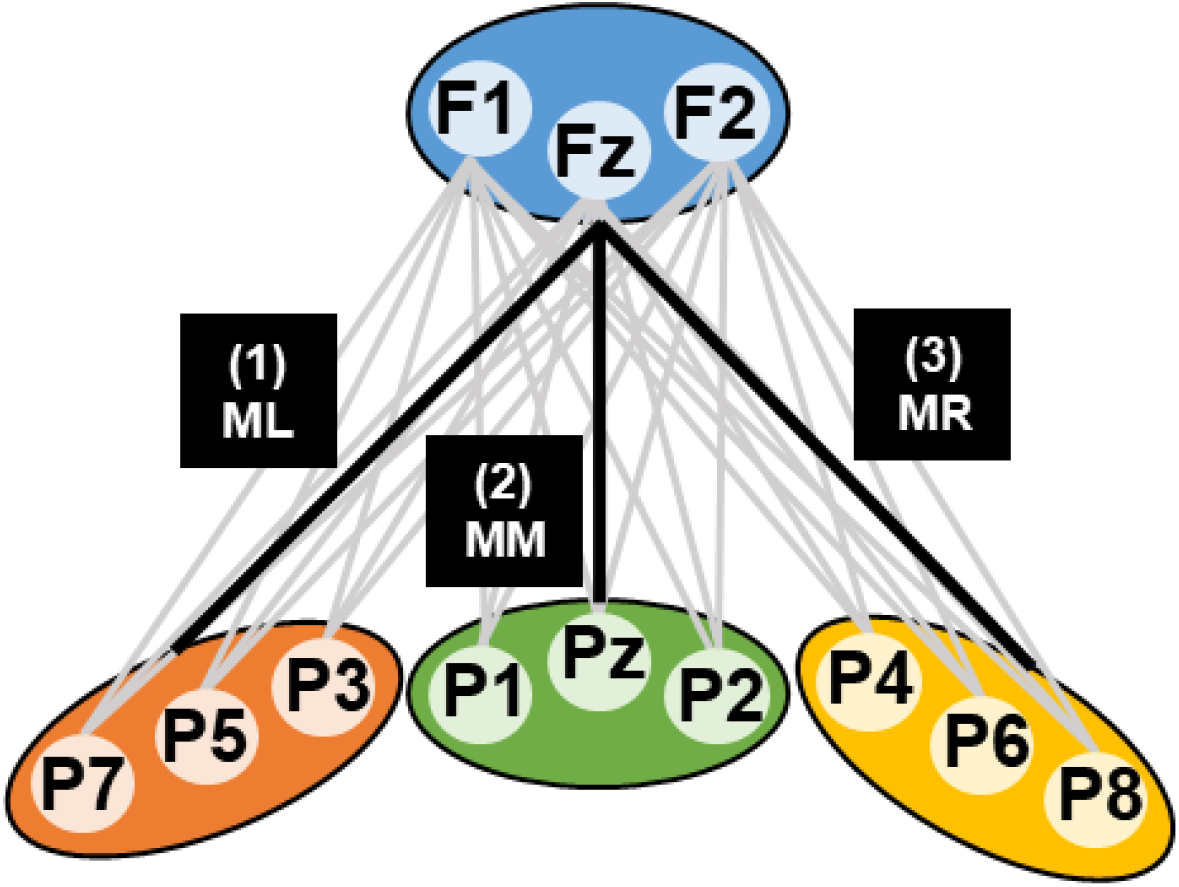
Graphical representation of the channels included in each region of interest. Grey lines indicate the 27 channel pairs that coherence was computed between. Black lines represent coherence averaged over electrode sets (ML: mid-frontal to left-parietal; MM: mid-frontal to mid-parietal; MR: mid-frontal to right-parietal).

Theta phase was computed for the whole epoch (0-1900ms) and then phase coherence was computed in the time-window of interest, 900-1500ms post-stimulus (the time-window ended at 1500ms to prevent inclusion of edge effects as per epoch definition). We calculated inter-site phase clustering (ISPC) [53]. ISPC over trials is a measure of the consistency of phase angles between two electrodes averaged over trials. For task-related data, ISPC-trials is an appropriate method given our analysis is hypothesis-driven (i.e., limited to the frontoparietal network) and not exploratory (more suited to weighted phase lag index) [53]. ISPC has been used previously in studies with similar methodology [52]. Phase angle time series for each channel were extracted by convolving the data with a complex Morlet wavelet (4 cycles) separately for frequencies 4Hz, 5Hz, 6Hz, and 7Hz. For each time point, the average vector length was calculated across trials to quantify ISPC-trials, defined as:

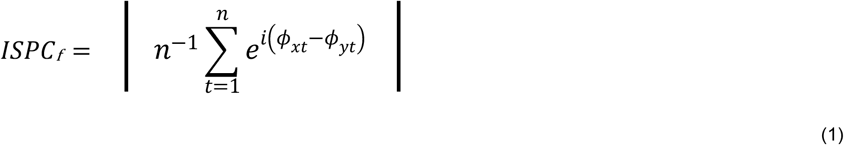

Where *n* represents the number of trials, and *ϕ* and *ϕ*_y_ are phase angles from channels *x* and *y* at frequency *f*. ISPC ranges from 0 (perfectly randomly distributed phases) to 1 (perfect phase-locking). For each channel pair ISPC-trials was calculated for each load (1-/2-back) and frequency (4-7Hz). The result was then averaged over the time-window of interest (900-1500ms post-stimulus), then over frequencies, and finally over channel sets. This resulted in one coherence value for each frontoparietal region pair/load combination (ML, MM, MR x 1-back, 2-back) for each participant.

### 2.5 Statistical analysis

Statistical analyses were conducted using SPSS version 25 [54]. The alpha level was set to 0.05. Visual inspection of Q-Q plots showed that, for each analysis, data were normally distributed. For each analysis of variance discussed below, Box and Whisker plots were inspected for extreme outliers. Values were considered extreme outliers if they fell outside of 3^rd^ quartile + 3*interquartile range and 1^st^ quartile – 3*interquartile range. Where extreme outliers were identified, sensitivity analyses were run. It can be assumed that there were no extreme outliers identified where sensitivity analysis is not reported.

In each frequency band a 2 (CON/NF1) x 2 (open/closed) analysis of variance (ANOVA) was run for the scalp averaged resting state data. ANOVAs were run separately for each frequency band as there is a known 1/f effect, whereby the means of low frequencies are larger than those of high frequencies [55]. As frequency bands are estimated independently, and therefore each ANOVA is performed on independent data, no correction for multiple comparisons was used. Moreover, to investigate whether resting state power follows the typical reactivity pattern observed in neurotypical populations [34], in each frequency band a paired *t*-test investigated power differences between eyes open and eyes closed resting state in the NF1 group. A 2 (CON/NF1) x 2 (open/closed) ANOVA was also used to analyse PAF.

In the n-back task, maintenance of items in working memory spans trials. We therefore used eyes-open resting state recordings as a baseline to investigate task-specific power modulation (i.e., change from rest). To achieve this, we divided task-related power by resting state power before log-transforming the data^6^ (equivalent to: log(task power) – log(resting state power)), which is referred to as *task-specific* power henceforth. In each frequency band a 2 (CON/NF1) x 2 (1-/2-back) ANOVA investigated scalp-averaged task-specific power. Consistent with the task-related power analyses we investigated task-specific theta phase coherence by adjusting for baseline (resting state). For comparability, eyes open resting state theta phase coherence was estimated using the same method as task-related phase coherence (note the ‘trials’ in ISPC-trials are arbitrary in resting state). Prior to statistical analysis resting state phase coherence was subtracted from task-related phase coherence. A 2 (CON/NF1) x 3 (ML/MM/MR) x 2 (1-/2-back) ANOVA using task-specific frontoparietal theta phase coherence was performed.

As existing research suggests a significant relationship between age and oscillatory activity in typically-developing children that may not be present in neurodevelopmental disorders [56], Pearson’s correlations were performed to investigate associations between EEG measures and age, followed by statistical significance testing of the difference in *r* between groups to determine whether the relationship between age and oscillatory activity was significantly different between groups. Moreover, as individuals with NF1 typically exhibit a lower average Intelligence Quotient (IQ) relative to typically-developing children [6,57], and there is suggestion that oscillatory activity might be a neural marker of cognitive function in neurodevelopmental disorders [56], Pearson’s correlations were performed to investigate associations between EEG measures and IQ, using Vineland ABC scores as a proxy. Again, this was followed by statistical significance testing of the difference in *r* between groups to determine whether the relationship between IQ and oscillatory activity was significantly different between groups. Finally, to assist interpretation of the oscillatory findings, Pearson’s correlations were performed to investigate associations between EEG measures and working memory performance on the adaptive auditory n-back task (which was conducted separately to the EEG session), and the difference in *r* between groups compared. To correct for multiple comparisons a 5% false discovery rate (FDR) [58] correction was applied to outcomes with *p*-values less than 0.05. FDR was applied to the set of EEG measures for each demographic/behavioural domain (i.e., five *p-*values).

## 3. Results

### 3.1 Behavioural

As expected, NF1^7^ performed significantly worse than CON on the parent-rated Vineland ABC (*t*_(29)_ = 3.573, *p*<.001) and adaptive auditory n-back task (mean n-back; *t*_(30)_ = 5.412, *p*<.001). Moreover, the NF1 group did not demonstrate any impairment in EEG n-back task performance (hits – false alarms (%); *F*_(1,30)_ = 0.094, *p*=.762, η_p_^2^=0.003) (see **Additional file 1** for detailed reporting).

### 3.2 Resting state power: higher delta and theta power in NF1

**Figure 2** displays spectral power as a continuous spectrum (top), and averaged in canonical frequency bands (middle), both averaged over all EEG channels, and as topographic maps (bottom) during **(a)** eyes open and **(b)** eyes closed resting state.

**Fig. 2.**
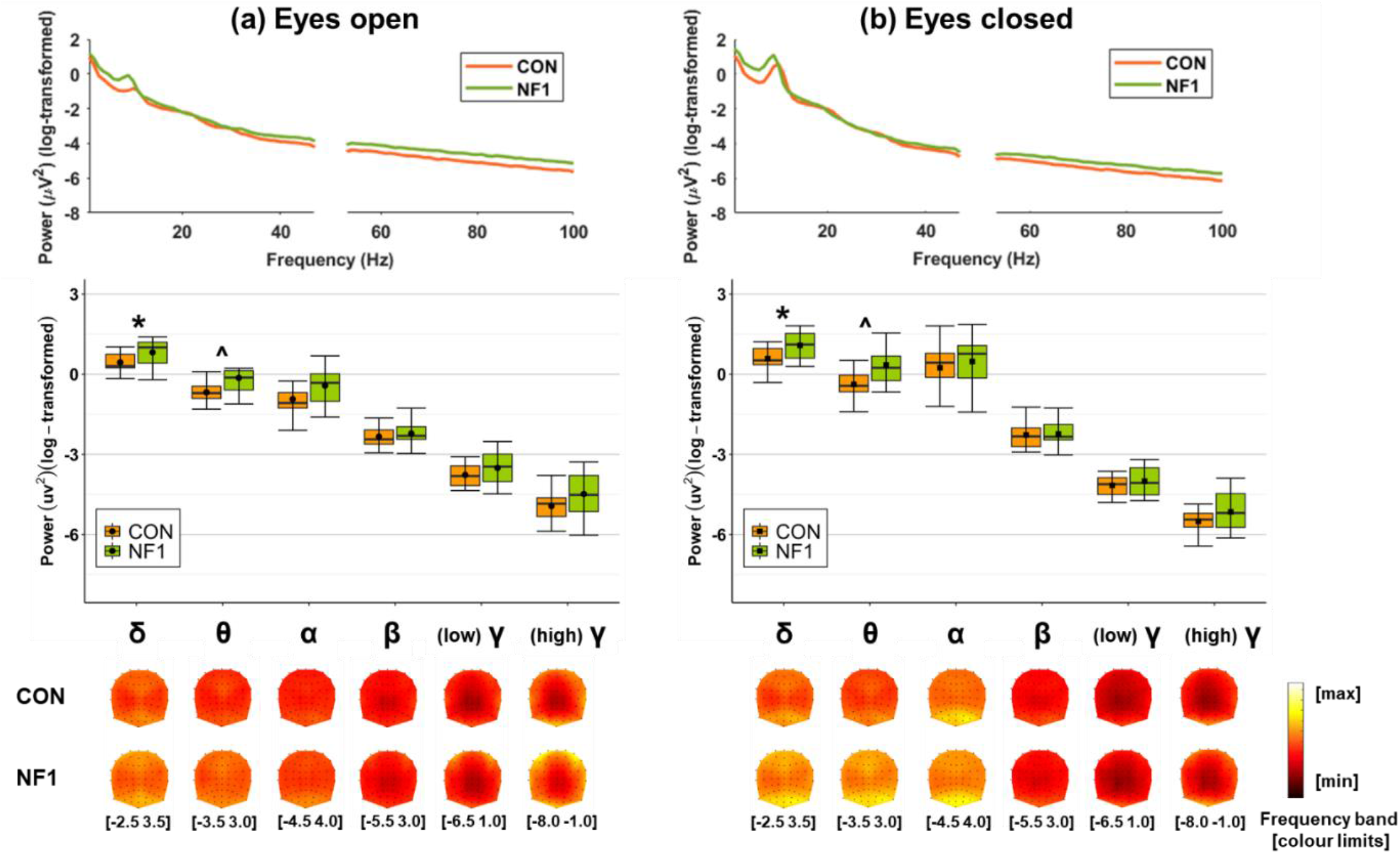
Grand-averaged log-transformed spectral power during rest with (a) eyes open and (b) eyes closed. Spectral power is shown as a continuous spectrum (top) and averaged in canonical frequency bands (middle), both averaged over all EEG channels, and as topographic maps (bottom). (Abbreviations: δ: delta, 1-3Hz; θ: theta, 4-7Hz; α: alpha, 8-11Hz; β: beta, 12-29Hz; (low) γ: low-gamma, 30-47Hz; (high) γ: high-gamma, 53-100Hz. Box plots: crossbars represent the median, dots represent the mean, upper and lower hinges correspond to the 1^st^ and 3^rd^ quartile, respectively, and whiskers represent the range of the data. * and ^ significant main effect of group in the delta and theta bands, respectively, collapsed over condition).

Visual inspection shows that spatial distributions were similar between CON and NF1 in all frequency bands, but with greater magnitudes in NF1 relative to CON. Moreover, greater magnitudes are seen during eyes closed relative to eyes open for delta, theta, and alpha, whilst the opposite pattern is observed for low-gamma and high-gamma. The difference in power between groups was significant for delta (*F*_(1,28)_ = 7.135, *p*=.012, η_p_^2^=.203) and theta (*F*_(1,28)_= 9.145, *p*=.005, η_p_^2^=.246), but non-significant for alpha, beta, low-gamma, and high-gamma **(Table 4, Figure 2)**.

**Table 4.**
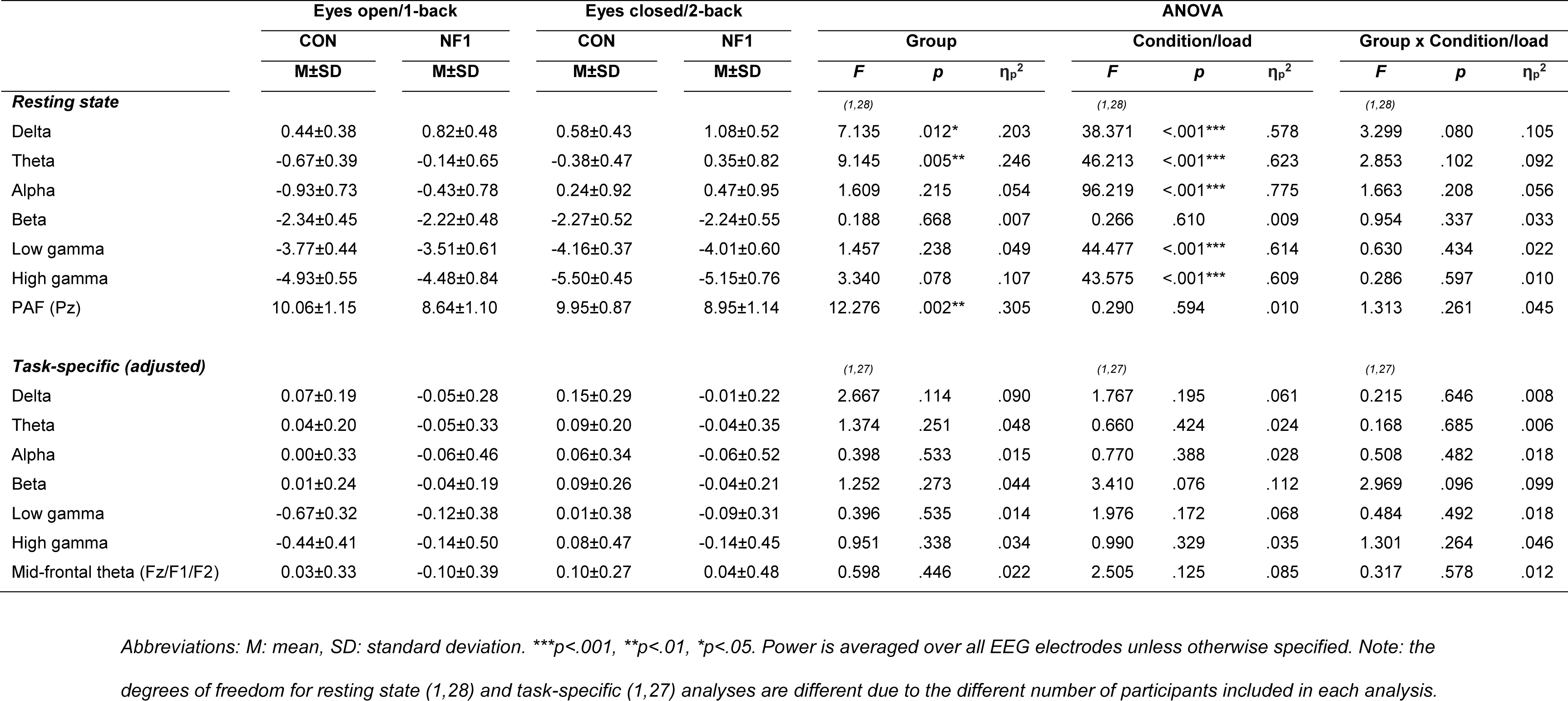
Power and PAF: descriptive and inferential statistics.

There were significant main effects of condition for delta, theta, alpha, low-gamma, and high-gamma, where power was significantly higher during eyes closed relative to eyes open for delta, theta, and alpha, but power was significantly higher during eyes open relative to eyes closed for low-gamma and high-gamma. The absence of significant group x condition interactions in these frequency bands suggests that modulation of amplitude of the oscillations did not differ between groups. Planned paired *t*-tests to examine oscillatory reactivity in the NF1 group showed that power was significantly higher during eyes closed relative to eyes open for delta (*t*_(13)_ = 5.004, *p*<.001, *d*=1.34), theta (*t*_(13)_ = 4.296, *p*=.001, *d*=1.15), and alpha (*t*_(13)_ = 5.291, *p*<.001, *d*=1.41), whilst power was significantly higher during eyes open relative to eyes closed for low-gamma (*t*_(13)_ = 4.457, *p*=.001, *d*=1.17) and high-gamma (*t*_(13)_ = 4.204, *p*=.001, *d*=1.12). There was no significant difference in power between eyes open and eyes closed for beta (*t*_(13)_ = 0.285, *p*=.780, *d*=0.08).

### 3.3 Peak alpha frequency: lower PAF in NF1

**Figure 3** illustrates the alpha range of grand average **(a)** eyes open and **(b)** eyes closed resting state after adjustment to reduce 1/f noise and the application of Gaussian smoothing. There was a significant difference in PAF between groups (*F*_(1,28)_ = 12.276, *p*=.002, η_p_^2^=.305), whereby compared to CON, NF1 showed a lower PAF **(Table 4)**. There was no main effect of condition, and no group x condition interaction.

**Fig. 3.**
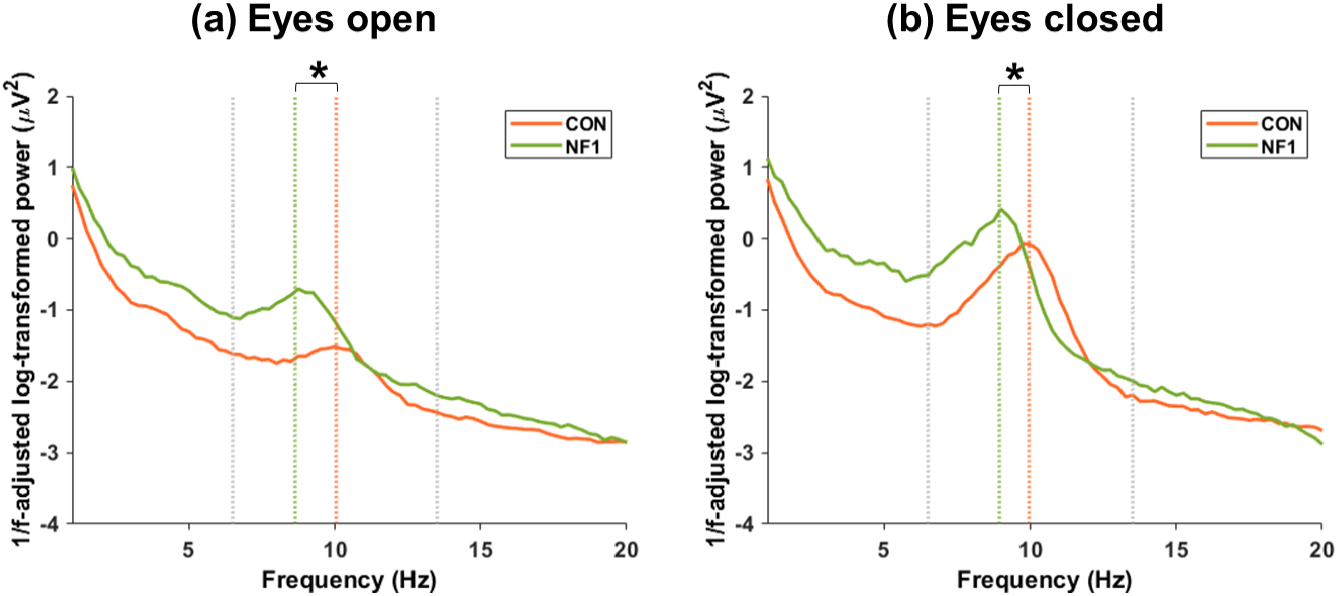
Grand-averaged 1/f-adjusted log-transformed spectral power during rest. (a) Eyes open and (b) eyes closed. (Grey dashed vertical lines at 6.5Hz and 13.5Hz represent the boundaries of the loose alpha range for PAF determination; orange and green dashed vertical lines represent the mean PAF for CON and NF1, respectively; * significant main effect of group collapsed over condition).

### 3.4 Task-related power: no group difference in task-specific power

**Figure 4** displays *unadjusted* spectral power as a continuous spectrum (top), and averaged in canonical frequency bands (middle), both averaged over all EEG channels, and as topographic maps (bottom) during **(a)** 1-back and **(b)** 2-back load levels.

**Fig. 4.**
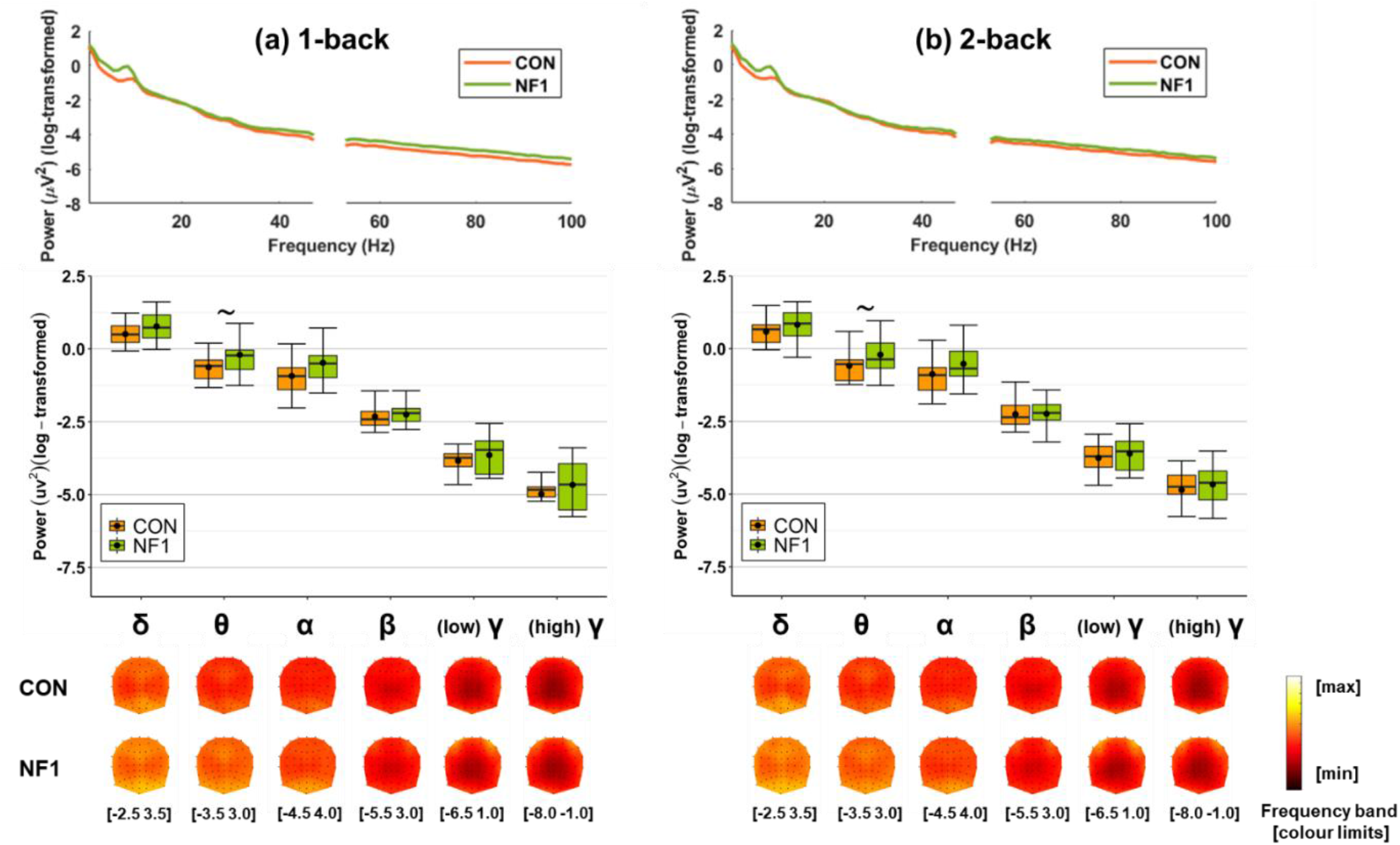
Grand-averaged log-transformed spectral *unadjusted* power during (a) 1-back and (b) 2-back loads. Spectral power is shown as a continuous spectrum (top) and averaged in canonical frequency bands (middle), both averaged over all EEG channels, and as topographic maps (bottom). (Abbreviations: δ: delta, 1-3Hz; θ: theta, 4-7Hz; α: alpha, 8-11Hz; β: beta, 12-29Hz; (low) γ: low-gamma, 30-47Hz; (high) γ: high-gamma, 53-100Hz. Box plots: crossbars represent the median, dots represent the mean, upper and lower hinges correspond to the 1^st^ and 3^rd^ quartile, respectively, and whiskers represent the range of the data. ∼ marginally significant main effect of group in the theta band, collapsed over load level).

Visual inspection shows that spatial distributions were similar between CON and NF1 in all frequency bands, but with greater magnitudes in NF1 relative to CON. This group difference in *unadjusted* spectral power was marginally significant for theta only (*F*_(1,29)_ = 4.092, *p*=.052, η_p_^2^=.124) (*unadjusted* power analyses are reported in **Additional file 2**). To investigate task-specific modulation we used task-related power adjusted for resting state eyes open power. Note, conclusions cannot be drawn about the absolute value of this difference (i.e., whether task-related is higher or lower than resting state activity) in the absence of a pre-stimulus baseline period in the n-back task. The 2 (CON/NF1) x 2 (1-/2-back) ANOVA^8^ showed no significant main effects or interactions in any frequency band **(Table 4)**. Thus, the marginal task-related difference in theta power disappeared when accounting for resting state theta power.

In line with the scalp-averaged *unadjusted* power, mid-frontal *unadjusted* theta power was numerically higher in NF1 than CON **(Figure 5)**^9^, though this group difference was non-significant (*F*_(1,29)_ = 2.850, *p*=.102, η_p_ ^2^=.089) **(Additional file 2)**. Similarly, the 2 (CON/NF1) x 2 (1-/2-back) ANOVA using task-specific *(adjusted)* theta power showed no significant main effects or interactions **(Table 4)**.

**Fig. 5.**
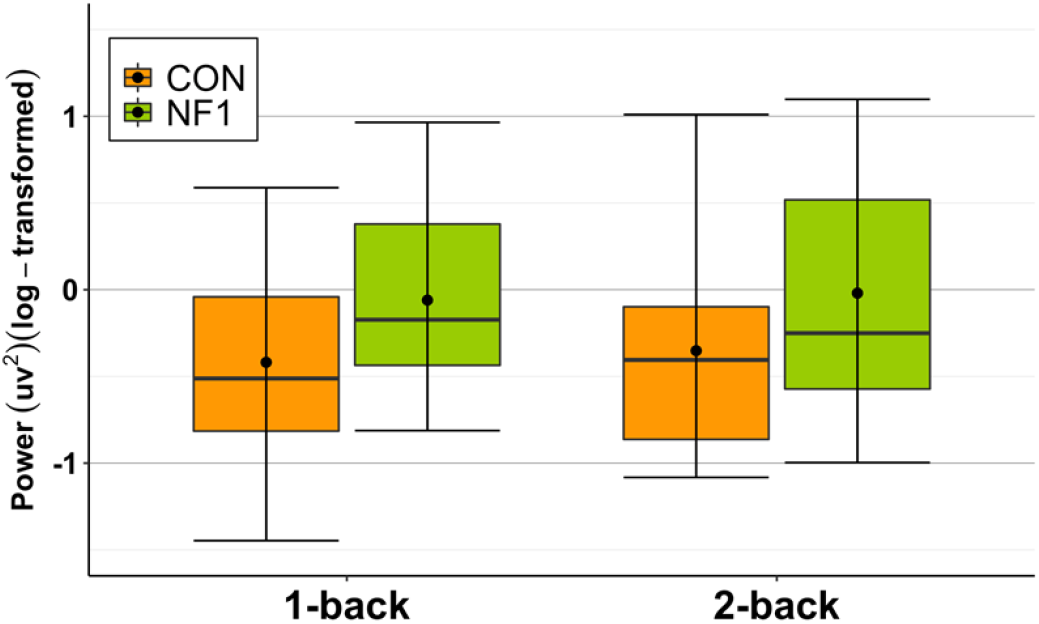
Grand-averaged log-transformed mid-frontal theta (4-7Hz) *unadjusted* power during (a) 1-back and (b) 2-back loads. (Box plots: crossbars represents the median, dots represents the mean, upper and lower hinges correspond to the 1^st^ and 3^rd^ quartile, respectively, and whiskers represent the range of the data).

### 3.5 Theta phase coherence: no group difference in task-specific phase coherence

Visual inspection of **Figure 6** shows that *unadjusted* frontoparietal theta phase coherence was numerically higher in NF1 relative to CON in all regions of the frontoparietal network during **(a)** 1-back and **(b)** 2-back loads. This group difference in *unadjusted* theta phase coherence was significantly different (*F*(1,29) = 4.852, *p*=.036, η_p_^2^=.143) **(Additional file 2)**. However, the 2 (CON/NF1) x 3 (ML/MM/MR) x 2 (1-/2-back) ANOVA^10^ using task-specific *(adjusted)* theta phase coherence showed no significant main effects or interactions involving the factor ‘group’ **(Table 5)**^11^. Again, conclusions cannot be drawn about the absolute value of this difference (i.e., whether task-related is higher or lower than resting state activity) in the absence of a pre-stimulus baseline period in the n-back task. Although not of primary interest, there was a significant region x load interaction (*F*_(2,54)_ = 5.023, *p*=.010, η_p_^2^=.157). There were no other significant main effects or interactions.

**Table 5.** Theta phase coherence *(adjusted)*: descriptive and inferential statistics.

| Descriptive statistics |  |  |  |  |
| --- | --- | --- | --- | --- |
| Region | Load | Group | M±SD |  |
| Mid-frontal –<br>left-parietal<br>(ML) | 1-back | CON | -0.014±0.028 |  |
|  |  | NF1 | -0.020±0.041 |  |
|  | 2-back | CON | -0.003±0.032 |  |
|  |  | NF1 | -0.014±0.055 |  |
| Mid-frontal –<br>mid-parietal<br>(MM) | 1-back | CON | -0.003±0.023 |  |
|  |  | NF1 | 0.006±0.049 |  |
|  | 2-back | CON | -0.003±0.034 |  |
|  |  | NF1 | -0.005±0.035 |  |
| Mid-frontal –<br>right-parietal<br>(MR) | 1-back | CON | -0.022±0.026 |  |
|  |  | NF1 | -0.131±0.021 |  |
|  | 2-back | CON | -0.120±0.032 |  |
|  |  | NF1 | -0.003±0.042 |  |
| ANOVA |  | *F*<sub>(1,27)/(2,54)</sub> | *p* | η<sup>2</sup><sub>p</sub> |
| Group |  | 0.028 | .868 | .001 |
| Region |  | 1.376 | .261 | .048 |
| Load |  | 1.505 | .231 | .053 |
| Group x region |  | 0.656 | .523 | .024 |
| Group x load |  | 0.580 | .453 | .021 |
| Region x load |  | 5.023 | .010* | .157 |
| Group x region x load |  | 0.503 | .608 | .018 |

Abbreviations: M: mean, SD: standard deviation. \* $p < .05$ . Degrees of freedom: (1,27), (2,54).

**Fig. 6.**
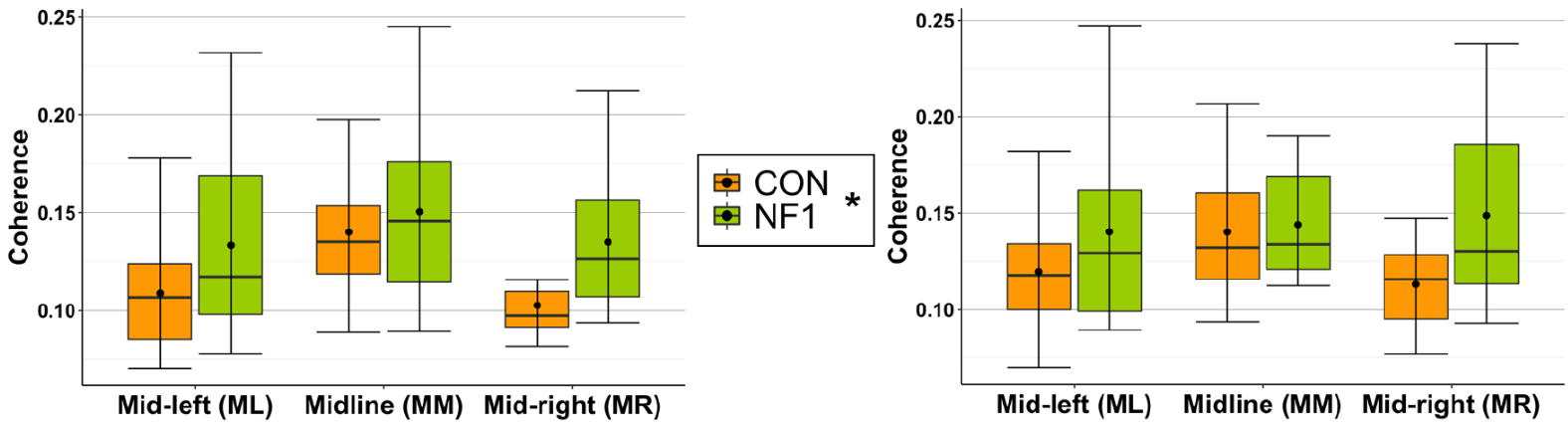
Box plots display *unadjusted* frontoparietal theta phase coherence during (a) 1-back and (b) 2-back loads. Mid-left (mid-frontal – left-parietal), midline (mid-frontal – mid-parietal), and mid-right (mid-frontal – right-parietal). (Crossbars represent the median, dots represent the mean, upper and lower hinges correspond to the 1^st^ and 3^rd^ quartile, respectively, and whiskers represent the range of the data. *significant main effect of group averaged over region and load).

### 3.6 Correlations between EEG measures and age/cognitive measures

Exploratory Pearson’s correlations were run separately for each group to relate individual differences in EEG oscillatory measures with age, overall cognitive function (ABC as a proxy for IQ), and working memory performance (adaptive auditory n-back) **(Table 6, see Additional file 4 for scatterplots)**. Correlations were run only for EEG measures that showed a significant group difference in the analyses reported above (i.e., delta and theta resting state power and PAF). For correlations with resting state power, as there was a significant main effect of condition for the delta and theta bands, correlations were run separately for eyes open and eyes closed power. There was no significant main effect of condition for PAF, so PAF was averaged over eyes open/closed prior to running correlations.

**Table 6.**
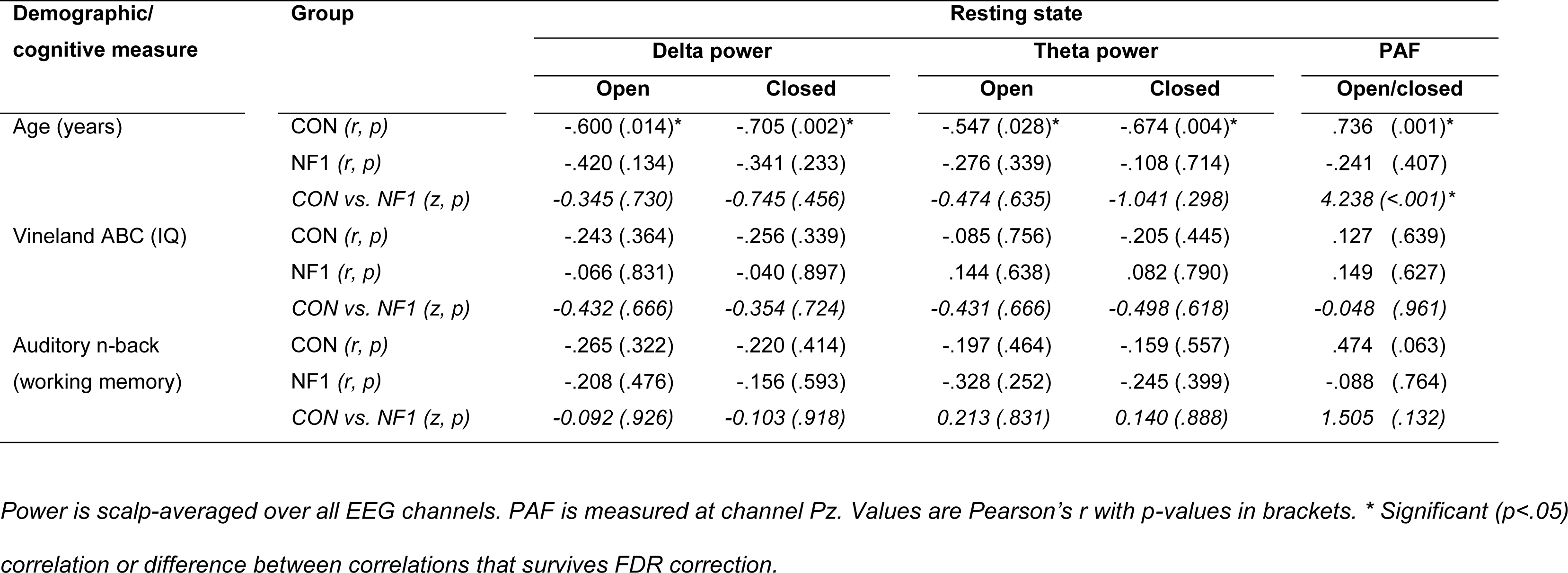
Correlations between EEG measures and age, Vineland ABC (proxy for IQ), and working memory.

For CON, four negative age-power correlations survived FDR correction: eyes open delta (*r*=-.600, *p*=.014), eyes closed delta (*r*=-.705, *p*=.002), eyes open theta (*r*=-.547, *p*=.028), and eyes closed theta (*r*=-.674, *p*=.004). The same correlations for NF1 were non-significant and group differences in these correlations were non-significant. Moreover, age showed a positive correlation with PAF for CON (*r*=.736, *p*=.001, survived FDR correction). The same correlation for NF1 was non-significant (*r*=-.241, *p*=.407) and the group difference in these correlations was significant (z=4.238, *p*<.001, survived FDR correction). Finally, there were no significant correlations between any of the EEG measures and overall cognitive function (ABC) or working memory performance.

## 4. Discussion

This study investigated oscillatory activity during both rest and performance of a working memory task in an adolescent sample with NF1 and age/sex-matched typically-developing controls. Relative to controls, NF1 showed higher resting state delta and theta power, and these differences were not modulated by eyes open/closed condition (no group x condition interactions were found). Resting state delta and theta power showed significant negative correlations with age in controls, but not in NF1. NF1 also showed lower PAF than controls, and the positive age-PAF correlation found in controls was not present in NF1 (and these correlations differed significantly between groups). In the working memory task, a marginal group difference in theta power was observed, but this effect disappeared when controlling for baseline (resting state) activity. Similarly, the significant group difference in frontoparietal theta phase coherence disappeared when values were adjusted for baseline (resting state). Together, these findings suggest that NF1 is characterised by aberrant resting state oscillatory activity and highlight the importance of accounting for resting state (baseline) differences when drawing conclusions about task-related differences in oscillatory activity between groups.

### 4.1 Resting state power

Resting state delta and theta power were significantly higher in NF1 than in typically-developing controls, in line with our hypothesis. This finding is consistent with, and builds on, previous reports in the NF1 cohort [17] and in other neurodevelopmental disorders [32]. For instance, Ribeiro et al. (2014) observed significantly higher theta power and a non-significant trend towards higher delta power in the NF1 cohort. Moreover, a review by Newson and Thiagarajan [32] of behaviourally-relevant frequency bands during resting state EEG in psychiatric disorders, including ADHD, reported that one of the most dominant abnormalities is increased power in slower frequencies.

Although the mechanisms underlying abnormally high slow wave activity in NF1 are not understood [17], previous studies using animal models of disrupted myelination have demonstrated that loss of myelin is associated with an increase in slow wave theta power [59]. The well-documented white matter microstructure abnormalities and myelin deficits in NF1 [60,61] could therefore account for the high slow wave resting state oscillatory activity observed in the current study.

Consistent with our prediction, direct tests of oscillatory power reactivity in the NF1 group showed that resting state power was significantly higher during eyes closed relative to eyes open in the delta, theta, and alpha bands, whilst the opposite pattern was observed in the gamma band. This demonstrates that resting state power reactivity follows the typical pattern observed in neurotypical populations [34] and builds on previous indirect suggestion (i.e., a non-significant group x condition interaction) reported in Ribeiro et al. [17].

Finally, delta and theta power showed significant negative correlations with age in typically-developing controls, consistent with existing literature showing that increasing age is associated with a reduction in slow wave resting state power throughout development [62,63]. The same correlations were non-significant for NF1, though there were no significant differences in correlations between groups. However, the relatively small sample size limits our ability to draw definitive conclusions about whether the relationship between age and oscillatory power is atypical in NF1.

### 4.2 Peak alpha frequency

PAF was significantly lower in the NF1 group relative to typically-developing controls. This builds on a non-significant trend towards a lower PAF observed in one previous study in NF1 [17] and is consistent with investigations of PAF in other neurodevelopmental disorders [55,64]. PAF is thought to reflect an index of cognitive preparedness [65], attentional processing [66], and memory ability [65,67]. Despite this, we did not observe a significant correlation between PAF and working memory ability using performance on an auditory n-back task in either the control or NF1 group. However, again, the relatively small sample size was not optimal to address this, and it is possible that significant associations may be found with a larger sample.

We observed a positive age-PAF correlation in typically-developing controls that was not present in NF1, and these correlations differed significantly between groups, suggesting that the relationship between PAF and age is disrupted in NF1. The age-PAF correlation in the control group is in accordance with existing literature in typically-developing children, where increased PAF is observed throughout childhood, stabilising at ∼10Hz during late adolescence/early adulthood [68]. This increase in PAF is thought to index neural network maturation [69,70] that facilitates improved and efficient connectivity [63,71]. Moreover, the absence of a significant PAF-age correlation in the NF1 group is consistent with previous research in other neurodevelopmental disorders that has reported the absence of a relationship between PAF and age [56]. It has been suggested that in neurodevelopmental disorders in which overall cognitive function (i.e., IQ) is disrupted and does not reliably map onto chronological age, PAF might instead be associated with IQ [56]. However, we were unable to provide direct support for this suggestion as the correlation between PAF and parent-rated functional ability measured by Vineland ABC was non-significant for NF1. Future studies measuring PAF (and other oscillatory measures) should consider including full-scale IQ testing using standardised measures and a non-NF1 developmentally delayed control group to disentangle generic effects of developmental delay and condition specific effects.

### 4.3 Task-related power and coherence

Task-related *(unadjusted)* theta power was significantly higher in the NF1 group relative to typically-developing controls, but this effect disappeared when controlling for baseline (resting state) activity. Likewise, the significant group difference in frontoparietal theta phase coherence (NF1>CON) disappeared when values were adjusted for baseline (resting state). The absence of a group difference in task-specific power and theta phase coherence may suggest that the NF1 population have a generally high level of oscillatory activity, particularly in low frequencies, which might explain the absence of a difference in task-specific modulation. These findings are inconsistent with our hypothesis that predicted aberrant WM-related theta power and phase coherence relative to controls based on existing research in other neurodevelopmental disorders [27,28,31,72]. However, this dissimilarity to observations in other neurodevelopmental disorders is not entirely surprising as, although neurodevelopmental disorders often exhibit overlapping cognitive impairments (i.e., strong clinical similarities), the underlying neurophysiology of these impairments is not always the same [14]. In sum, the current study suggests that task-related oscillatory activity might not be a useful EEG marker of working memory impairment in NF1. Instead, a better EEG marker of working memory impairment in NF1 might be the ERP P3b component, which has been found to differ in latency and topographic distribution in NF1 relative to typically-developing controls [16].

### 4.4 Comorbid ADHD

Individuals with comorbid NF1 and ADHD exhibit a more severe cognitive deficit than individuals with NF1 but without ADHD [73]. Therefore, in line with Ribeiro et al. [17], we ran sensitivity analyses **(Additional file 5)** to determine whether the significant group differences observed remained after removing participants with comorbid ADHD. Four of the sixteen participants in the NF1 group had an ADHD diagnosis, reducing the NF1 group size to 12. Following sensitivity analyses, the significant group difference in resting state theta power (*p*=.012) and PAF (*p*=.005) remained. However, the group difference in resting state delta power was only marginally significant (*p*=.066). Though, these sensitivity analyses should be interpreted with caution owing to the reduced, and subsequently small, group size and should act as a starting point for future studies using larger samples [17]. Like Ribeiro et al. [17] we were unable to compare individuals with comorbid ADHD and NF1 to controls due to the very small number (n=4) of participants with this comorbidity. Thus, future studies comparing these populations would facilitate an understanding of whether this comorbidity results in a different EEG profile to the one we found.

### 4.5 Strengths, limitations, and future directions

This study has informed the characterisation of NF1 and potential targets (e.g., abnormally high slow wave power) for (non-) pharmacological interventions targeting NF1. However, this contribution must be considered in light of several limitations. Our sample size was comparable to existing neuroimaging studies using NF1 samples [13,14,15,17,18]; however, post-hoc power analysis of Ribeiro et al.’s [17] resting state theta effect suggests that slightly larger sample sizes would be needed to achieve adequate statistical power (effect size of d=0.89, one-tailed t-test, 80% power requires n=17 per group calculated using G*Power [74]). Further, we were likely underpowered for the examination of associations between oscillatory features and age/cognitive measures. Future studies with larger sample sizes would help to draw more definitive conclusions regarding individual differences in oscillatory abnormalities in NF1. A second limitation of this study is that, whilst we investigated group differences in single frequency bands, it may be beneficial to examine the relationships between different frequency bands during working memory performance to further understand the basis of working memory deficits in NF1. For instance, theta-gamma phase-amplitude coupling is a neural marker associated with working memory performance [20] and has provided a neural marker of poor working memory in other clinical disorders (e.g., schizophrenia, Alzheimer’s Disease, and Mild Cognitive Impairment [75,76]).

## 5. Conclusions

This study investigated oscillatory activity both at rest and during performance of a working memory task. We found that adolescents with NF1 display aberrant oscillatory activity relative to typically-developing controls during rest. Specifically, NF1 is characterised by excessive resting state oscillatory activity, particularly at lower frequencies, and a lower peak alpha frequency. Moreover, we found that apparent group differences in working memory task-related oscillatory power and frontoparietal coherence disappeared when accounting for baseline levels from resting state. These findings provide insights that can inform the characterisation of NF1, as well as the design of (non-) pharmacological interventions targeting NF1, and also highlight important avenues for future research.

## Data Availability

The datasets used and/or analysed during the current study are available from the corresponding author on reasonable request.

## 6. Abbreviations

ADHD: Attention-deficit hyperactivity disorder
ASD: Autism spectrum disorder
CON: Control
EEG: Electroencephalography
ERP: Event-Related Potential
fMRI: Functional Magnetic Resonance Imaging
ICA: Independent Components Analysis
IQ: Intelligence Quotient
ISPC: Inter-site phase clustering
M: Mean
ML: Mid-frontal – left-parietal
MM: Mid-frontal – mid-parietal
MR: Mid-frontal – right-parietal
NF1: Neurofibromatosis Type 1
PAF: Peak alpha frequency
SD: Standard deviation
VABS-II: Vineland Adaptive Behaviour Scale
Vineland ABC: Vineland Adaptive Behaviour Composite

## 7. Declarations

### Ethics approval and consent to participate

This study received ethical approval from the Greater Manchester West Research Ethics Committee (17/NW/0364) and was conducted in accordance with the Declaration of Helsinki. Parents/guardians gave oral and written consent, and adolescents assent (where developmentally appropriate), prior to participation.

### Consent for publication

Not applicable.

### Competing interests

The authors declare that they have no competing interests.

### Funding

Data collection was funded by the Newlife Charity for Disabled Children. The analysis and write-up was conducted as part of a PhD funded by the Economic and Social Research Council (ESRC), awarded to Miss Samantha Booth. The funders had no role in the study design, data collection, data analysis, statistical analysis, interpretation of the data, writing of the paper, or decision regarding when to submit the publication.

### Authors’ contributions

SB, SG, JG, GP, and JT conceptualised and designed the study. SG was responsible for participant recruitment. SG, GP, and JT were responsible for data collection. SB analysed the data and wrote the original manuscript draft. SB, SG, LB, GP, and JT contributed towards interpretation of the data. All authors contributed towards reviewing and editing the manuscript. All authors read and approved the final manuscript.

## Acknowledgements

We would like to thank the participants and their families for their participation, and Emily Pye, JeYoung Jung, and Hemavathy Ramalingham for their roles in data collection.

## Additional files

### Additional file 1

- **Format:** Word document (.pdf)
- **Title:** Sub-sample demographic and behavioural information
- **Description: Table 1**. Descriptive and inferential statistics for age, sex, Vineland ABC scores, and auditory n-back performance. **Table 2**. Descriptive and inferential statistics for EEG visual n-back task performance (hits % – false alarms %).

### Additional file 2

- **Format:** Word document (.pdf)
- **Title:** Statistical analysis of task-related *unadjusted* power and theta coherence
- **Description: Table 1**. Power *(unadjusted)*: descriptive and inferential statistics. **Table 2**. Theta phase coherence *(unadjusted)*: descriptive and inferential statistics.

### Additional file 3

- **Format:** Word document (.pdf)
- **Title:** Resting state analyses for mid-frontal theta power and theta phase coherence
- **Description: Table 1**. Resting state (eyes open) mid-frontal theta power: descriptive and inferential statistics. **Table 2**. Resting state (eyes open) theta phase coherence: descriptive and inferential statistics.

### Additional file 4

- **Format:** Word document (.pdf)
- **Title:** Scatterplots
- **Description: Fig. 1**. Scatterplots between EEG measures and age. **Fig. 2**. Scatterplots between EEG measures and Vineland ABC scores (IQ). **Fig. 3**. Scatterplots between EEG measures and auditory n-back performance (working memory).

### Additional file 5

- **Format:** Word document (.pdf)
- **Title:** Sensitivity analysis
- **Description: Table 1**. Sensitivity analyses outcomes.

## Footnotes

1 See Section 4.5 for a consideration of power.

2 This remained the case for the sub-group of participants included in the resting state (CON/NF1: n=16/14) and task-specific (CON/NF1: n=16/13) analyses **(Additional file 1)**.

3 Parents completed the pen-and-paper version of the Vineland ABC.

4 Note, the use of ICA for artefact signal removal does not pose a problem for phase-based analyses such as coherence [45]. ICA is an instantaneous spatial filtering method, and as such it does not distort the phases of the underlying signals. There may, however, be *apparent* changes in phase of *channel* data after removing artefact components since channels contain weighted sums of underlying neural and artefact signals (some of which have been removed). In fact, after artefact removal, channel data should be a purer measure of neural source data, including the oscillations of interest here.

5 For task-related data, the number of trials remaining reflects the number of trials after incorrect responses were removed.

6 To calculate task-specific power and phase coherence participants needed to be eligible for both resting state and task-related analyses. This reduced the sample size from N=31 (CON/NF1: n=16/15) to N=29 (CON/NF1: n=16/13).

7 These findings remained the same when the analyses were run on the sample eligible for inclusion (see Section 2.4) in the resting state (N=30), task-related (N=31), and task-specific analyses (N=29) reported the current paper **(Additional file 1)**.

8 One extreme outlier was identified in the NF1 group (beta 2-back) from inspection of Box and Whisker plots. After removing this participant from the beta ANOVA, the findings stayed the same (i.e., no significant main effects or interactions).

9 Resting state mid-frontal theta power descriptive and inferential statistics are reported in **Additional file 3**.

10 Two extreme outliers were identified in the NF1 group (ML 2-back) from inspection of Box and Whisker plots. After removing these outliers, the findings remained the same (i.e., a significant region x load interaction but no other significant main effects or interactions).

11 Resting state theta phase coherence descriptive and inferential statistics are reported in **Additional file 3**.

## Notes

### Competing Interest Statement

The authors have declared no competing interest.

### Author Declarations

This study received ethical approval from the Greater Manchester West Research Ethics Committee (17/NW/0364) and was conducted in accordance with the Declaration of Helsinki. Trial registration: ClinicalTrials.gov identifier: NCT03310996 (first posted: October 16 2017). Parents/guardians gave oral and written consent, and adolescents assent (where developmentally appropriate), prior to participation.

