## Additional File 1 for "Aberrant oscillatory activity in Neurofibromatosis Type 1: An EEG study of resting state and working memory"

### Sub-sample demographic and behavioural information

**Table 1.** Descriptive and inferential statistics for age, sex, Vineland ABC scores, and auditory n-back performance.

| | Data | Descriptives: CON | Descriptives: NF1 | *t*-test or Chi-square test |
|---|---|---|---|---|
| **Demographics** | | | | |
| Age (M±SD (range)) | Resting state | 13.34±1.61 (11.33-16.92) | 13.09±1.69 (11.25-16.58) | $t_{(28)} = 0.412$, $p$=.683 |
| | Task-related (unadjusted) | 13.34±1.61 (11.33-16.92) | 13.05±1.72 (11.25-16.58) | $t_{(29)} = 0.483$, $p$=.633 |
| | Task-specific (adjusted) | 13.34±1.61 (11.33-16.92) | 13.12±1.76 (11.25-16.58) | $t_{(27)} = 0.346$, $p$=.732 |
| Sex (male/female) | Resting state | 9/7 | 8/6 | $\chi2 = 0.002$, $p$=.961 |
| | Task-related (unadjusted) | 9/7 | 8/7 | $\chi2 = 0.027$, $p$=.870 |
| | Task-specific (adjusted) | 9/7 | 7/6 | $\chi2 = 0.017$, $p$=.897 |
| **Behavioural** | | | | |
| Vineland ABC[1] | Resting state | 103.56±15.35 | 81.85±14.31 | $t_{(27)} = 3.904$, $p$=.001 |
| | Task-related (unadjusted) | 103.56±15.35 | 85.00±15.36 | $t_{(28)} = 3.303$, $p$=.003 |
| | Task-specific (adjusted) | 103.56±15.35 | 84.00±12.56 | $t_{(26)} = 3.598$, $p$=.001 |
| Auditory n-back (mean n-back) | Resting state | 2.64±0.38 | 1.98±0.36 | $t_{(28)} = 4.872$, $p$=<.001 |
| | Task-related (unadjusted) | 2.64±0.38 | 1.91±0.37 | $t_{(29)} = 5.461$, $p$=<.001 |
| | Task-specific (adjusted) | 2.64±0.38 | 1.96±0.37 | $t_{(27)} = 4.884$, $p$=<.001 |

*Abbreviations: M: mean, SD: standard deviation.*

[1] One participant in the NF1 group did not have an ABC score.

**Table 2.** Descriptive and inferential statistics for EEG visual n-back task performance (hits % – false alarms %).

| Data | 1-back | | 2-back | | ANOVA | | | | | | | | |
|---|---|---|---|---|---|---|---|---|---|---|---|---|---|
| | CON | NF1 | CON | NF1 | Group | | | Load | | | Group x load | | |
| | M±SD (%) | M±SD (%) | M±SD (%) | M±SD (%) | *F* | *p* | $\eta_p^2$ | *F* | *p* | $\eta_p^2$ | *F* | *p* | $\eta_p^2$ |
| Resting-state | 83.81±17.50 | 83.56±16.18 | 64.69±22.21 | 61.19±21.46 | 0.055 | .816 | .002 | 36.097 | <.001 | .563 | 0.057 | .812 | .002 |
| Task-related (unadjusted) | 83.81±17.50 | 84.00±16.65 | 64.69±22.21 | 61.20±22.22 | 0.068 | .796 | .002 | 41.167 | <.001 | .587 | 0.316 | .578 | .011 |
| Task-specific (adjusted) | 83.81±17.50 | 83.54±17.92 | 64.69±22.21 | 62.46±23.37 | 0.034 | .855 | .001 | 34.136 | <.001 | .558 | 0.080 | .779 | .003 |

*Abbreviations: M: mean, SD: standard deviation.*
