## Additional File 2 for "Aberrant oscillatory activity in Neurofibromatosis Type 1: An EEG study of resting state and working memory"

### Statistical analysis of task-related *unadjusted* power and theta coherence

**Table 1.** Power *(unadjusted)*: descriptive and inferential statistics.

| | 1-back | | 2-back | | ANOVA | | | | | | | | |
|---|---|---|---|---|---|---|---|---|---|---|---|---|---|
| | CON | NF1 | CON | NF1 | Group | | | Load | | | Group x load | | |
| | M±SD | M±SD | M±SD | M±SD | $F_{(1,29)}$ | $p$ | $\eta_p^2$ | $F_{(1,29)}$ | $p$ | $\eta_p^2$ | $F_{(1,29)}$ | $p$ | $\eta_p^2$ |
| Delta | 0.51±0.39 | 0.77±0.53 | 0.59±0.48 | 0.82±0.59 | 2.051 | .163 | .066 | 2.236 | .146 | .072 | 0.122 | .729 | .004 |
| Theta | -0.63±0.47 | -0.20±0.64 | -0.59±0.51 | -0.20±0.64 | 4.092 | .052 | .124 | 0.374 | .545 | .013 | 0.465 | .501 | .016 |
| Alpha | -0.93±0.74 | -0.48±0.73 | -0.87±0.79 | -0.52±0.73 | 2.296 | .141 | .073 | 0.053 | .819 | .002 | 1.675 | .206 | .055 |
| Beta | -2.32±0.45 | -2.25±0.50 | -2.25±0.50 | -2.24±0.48 | 0.058 | .812 | .002 | 4.136 | .051 | .125 | 1.836 | .186 | .060 |
| Low gamma | -3.84±0.41 | -3.64±0.65 | -3.76±0.51 | -3.61±0.56 | 0.848 | .365 | .028 | 2.005 | .167 | .065 | 0.358 | .554 | .012 |
| High gamma | -4.98±0.53 | -4.67±0.84 | -4.86±0.66 | -4.67±0.68 | 1.117 | .299 | .037 | 0.814 | .374 | .027 | 0.856 | .363 | .029 |
| Mid-frontal theta (Fz/F1/F2) | -0.42±0.59 | -0.06±0.57 | -0.35±0.57 | -0.02±0.67 | 2.850 | .102 | .089 | 0.559 | .461 | .019 | 0.034 | .855 | .001 |

*Abbreviations: M: mean, SD: standard deviation. Power is averaged over all EEG electrodes unless otherwise specified. Degrees of freedom $_{(1,28)}$.*

**Table 2.** Theta phase coherence *(unadjusted)*: descriptive and inferential statistics.

| Descriptive statistics | | | | |
|---|---|---|---|---|
| **Region** | **Load** | **Group** | **M±SD** | |
| Mid-frontal – left-parietal (ML) | 1-back | CON | 0.109±0.028 | |
| | | NF1 | 0.133±0.049 | |
| | 2-back | CON | 0.120±0.030 | |
| | | NF1 | 0.140±0.048 | |
| Mid-frontal – mid-parietal (MM) | 1-back | CON | 0.140±0.037 | |
| | | NF1 | 0.150±0.047 | |
| | 2-back | CON | 0.140±0.034 | |
| | | NF1 | 0.144±0.028 | |
| Mid-frontal – right-parietal (MR) | 1-back | CON | 0.103±0.017 | |
| | | NF1 | 0.135±0.035 | |
| | 2-back | CON | 0.113±0.022 | |
| | | NF1 | 0.149±0.044 | |
| **ANOVA** | | $F_{(1,29)/(2,58)}$ | $p$ | $\eta_p^2$ |
| Group | | 4.852 | .036* | .143 |
| Region | | 5.736 | .005** | .165 |
| Load | | 2.865 | .101 | .090 |
| Group x region | | 2.309 | .108 | .074 |
| Group x load | | 0.114 | .739 | .004 |
| Region x load | | 4.700 | .013* | .139 |
| Group x region x load | | 0.473 | .625 | .016 |

*Abbreviations: M: mean, SD: standard deviation. ***p<.001, **p<.01, *p<.05. Degrees of freedom: $_{(1,29), (2,58)}$.*
