## Additional File 3 for "Aberrant oscillatory activity in Neurofibromatosis Type 1: An EEG study of resting state and working memory"

**Resting state analyses for mid-frontal theta power and theta phase coherence**

The NF1 group showed significantly higher mid-frontal theta power **(Table 1)** and frontoparietal theta phase coherence **(Table 2)** during resting state (eyes open) relative to controls.

**Table 1.** Resting state (eyes open) mid-frontal theta power: descriptive and inferential statistics.

| | **Descriptives** | | ***t*-test** | |
|---|---|---|---|---|
| **Group** | **M±SD** | $t_{(28)}$ | $p$ | **d** |
| CON | -0.45±0.40 | 2.619 | .014* | 0.93 |
| NF1 | 0.02±0.58 | | | |

*Abbreviations: M: mean, SD: standard deviation. *p<.05. Degrees of freedom:* $_{(28)}$.

**Table 2.** Resting state (eyes open) theta phase coherence: descriptive and inferential statistics.

| **Descriptive statistics** | | | |
|---|---|---|---|
| **Region** | **Group** | **M±SD** | |
| Mid-frontal – left-parietal (ML) | CON | 0.122±0.021 | |
| | NF1 | 0.159±0.062 | |
| Mid-frontal – mid-parietal (MM) | CON | 0.143±0.040 | |
| | NF1 | 0.153±0.038 | |
| Mid-frontal – right-parietal (MR) | CON | 0.125±0.032 | |
| | NF1 | 0.155±0.044 | |
| **ANOVA** | $F_{(1,28)}$ | $p$ | $\eta_p^2$ |
| Group[1] | 4.329 | .047* | .134 |
| Region | 0.725 | .489 | .025 |
| Group x region | 1.591 | .213 | .054 |

*Abbreviations: M: mean, SD: standard deviation. *p<.05. Degrees of freedom:* $_{(1,28)}$.

[1] One extreme outlier was identified in the CON group (ML) from inspection of a Box and Whisker plot. After removing this outlier, the findings stayed the same (i.e., a significant main effect of group, but no significant main effect of region or group x region interaction).
