## Additional File 4 for "Aberrant oscillatory activity in Neurofibromatosis Type 1: An EEG study of resting state and working memory"

**Scatterplots**

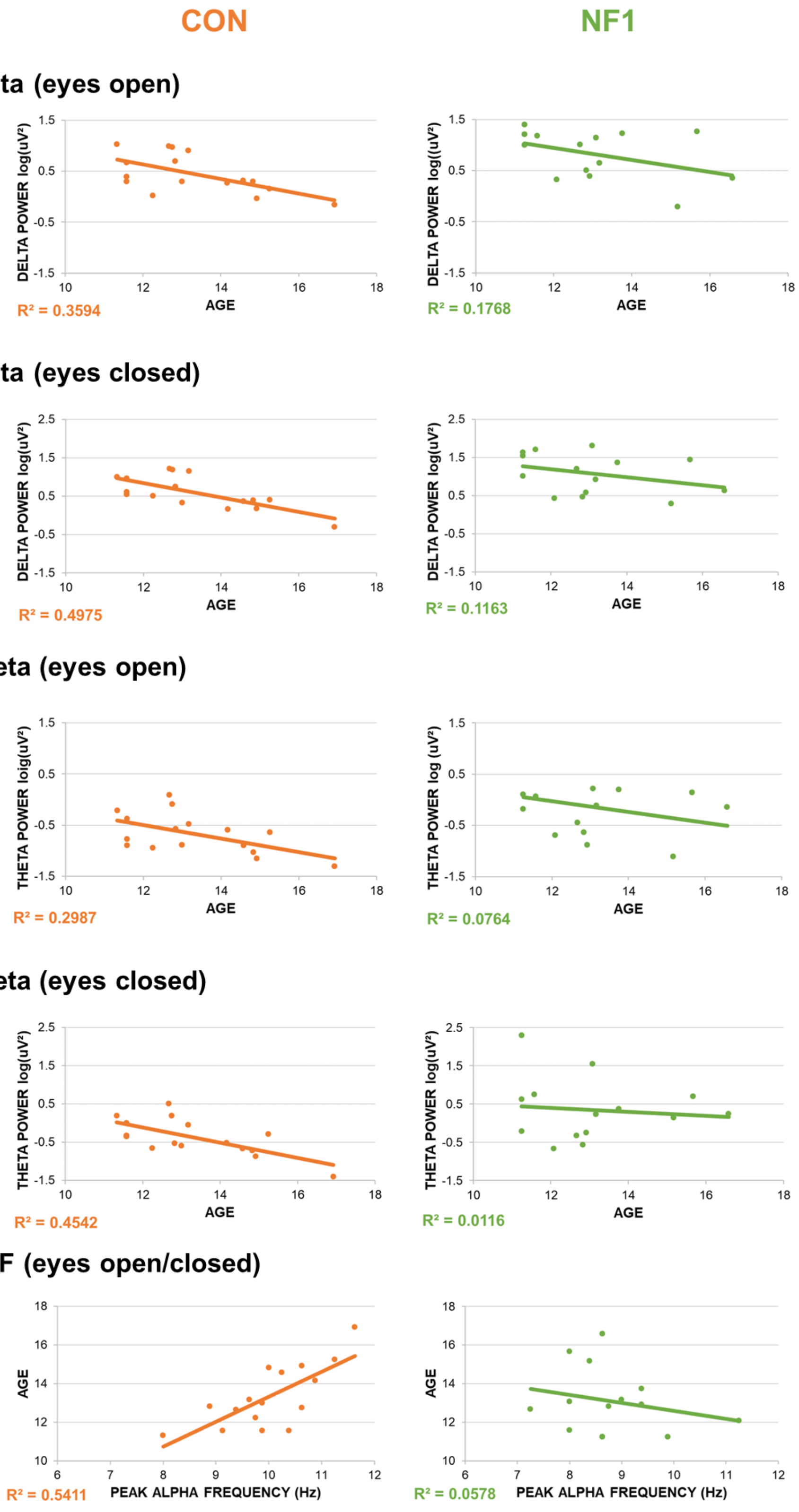


**Fig. 1.** Scatterplots between EEG measures and age.

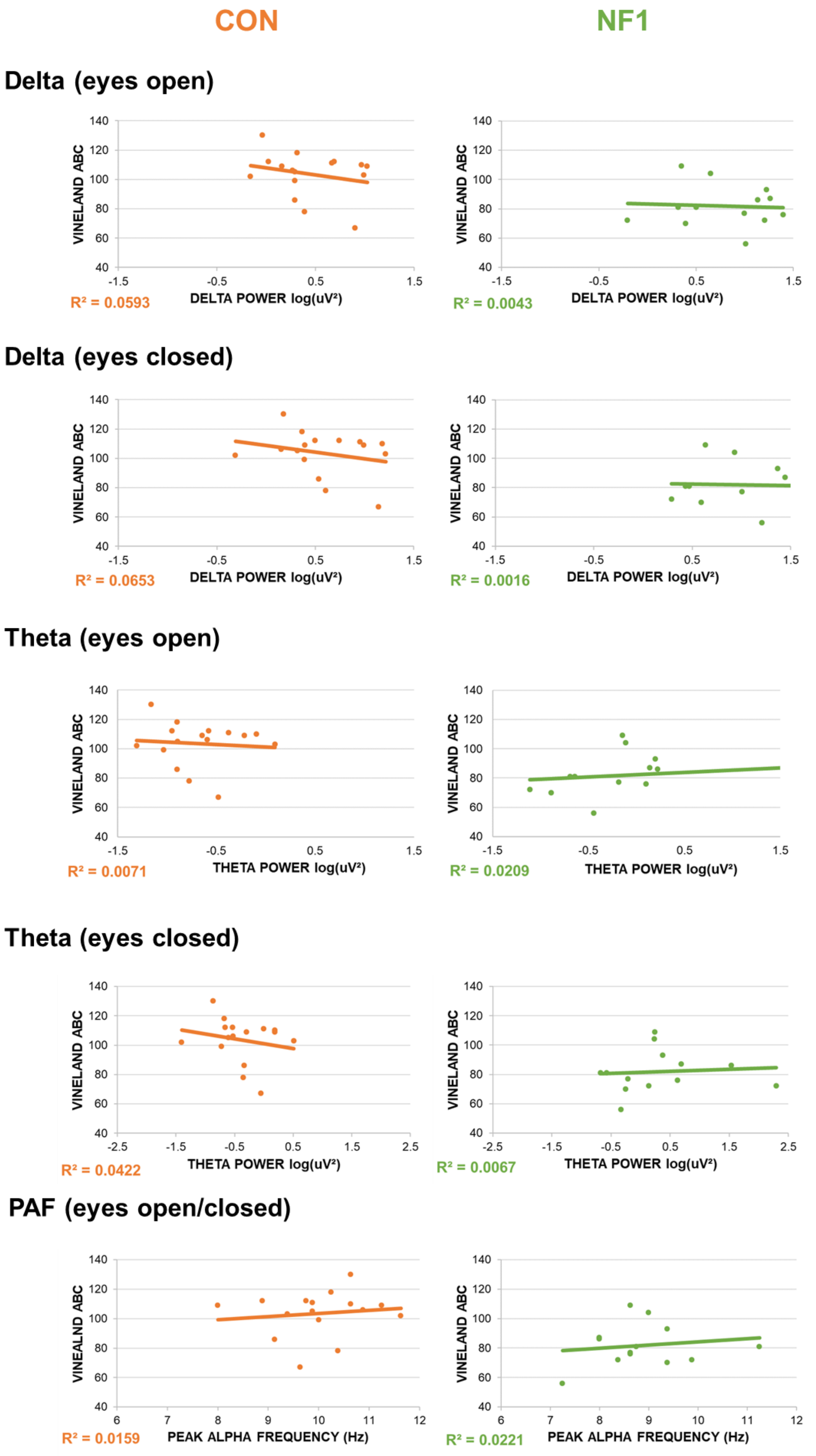


**Fig. 2.** Scatterplots between EEG measures and Vineland ABC scores (IQ).

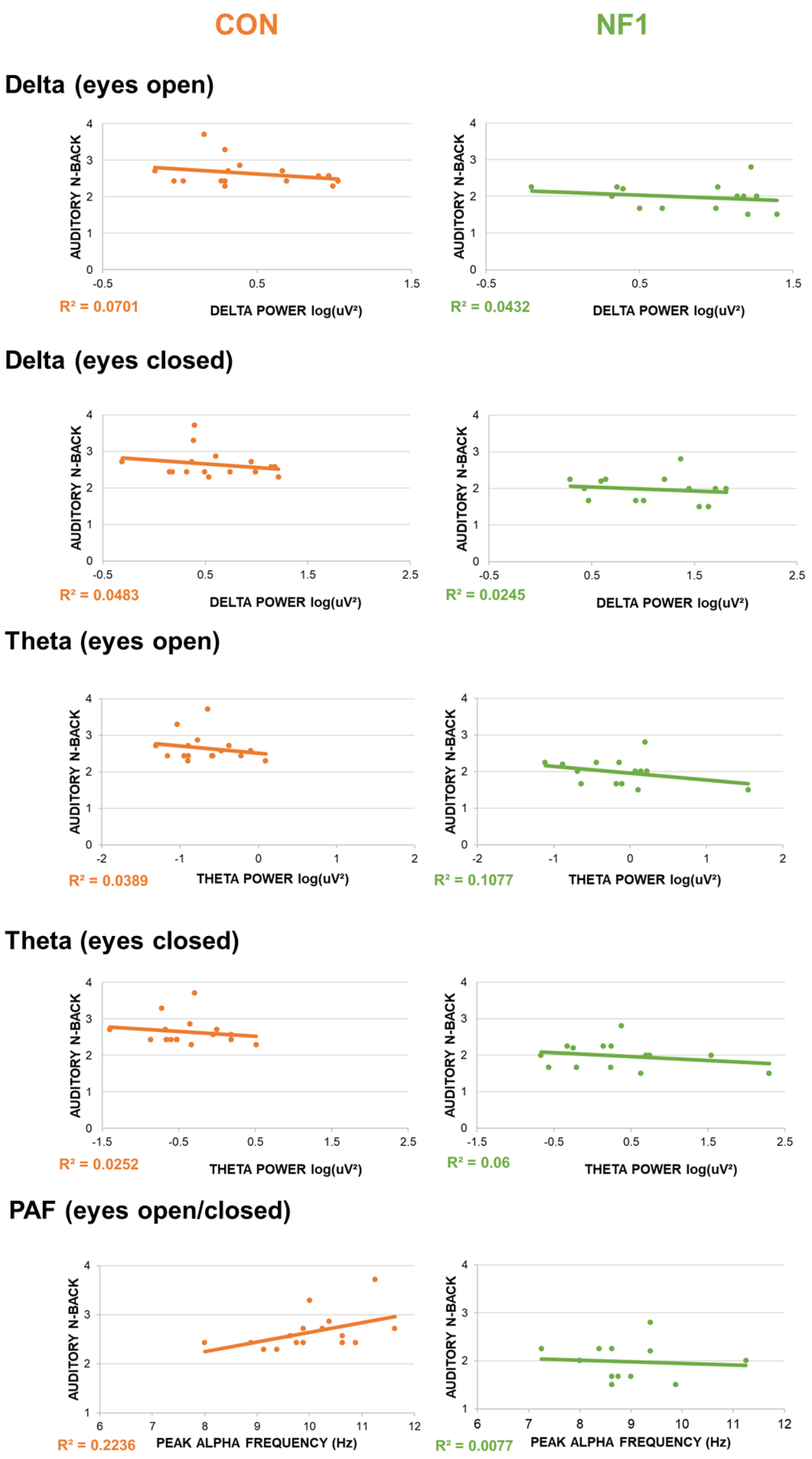


**Fig. 3.** Scatterplots between EEG measures and auditory n-back performance (working memory).
