## Additional File 5 for "Aberrant oscillatory activity in Neurofibromatosis Type 1: An EEG study of resting state and working memory"

### Sensitivity analyses

Existing research has shown that individuals with comorbid NF1 and ADHD exhibit a more severe cognitive deficit [1]. Therefore, in line with Ribeiro et al. (2014), we ran sensitivity analyses whereby participants with comorbid ADHD were removed from each analysis to determine whether the significant group differences we observed between NF1 and CON remained. Four of the sixteen participants in the NF1 group had an ADHD diagnosis. This means that the group size for the NF1 sample reduced to 12 once they were removed from the dataset. Therefore, any findings should be interpreted with caution owing to the small group size the analyses were performed on **(Table 1).**

**Table 1.** Sensitivity analyses outcomes.

| | ***Complete data set as reported in the main text*** | ***Participants with ADHD removed from the analysis*** |
|---|---|---|
| Resting state power | Higher delta (*p*=.012) and theta (*p*=.005) power in NF1 compared to CON. | Group difference in delta power is marginally significant (*p*=.066). Group difference in theta power remains significant (*p*=.012). |
| Peak alpha frequency (PAF) | Lower PAF in NF1 compared to CON (*p*=.002). | Group difference remains significant (*p*=.005). |
